# Stability of SARS-CoV-2 on environmental surfaces and in human excreta

**DOI:** 10.1101/2020.05.07.20094805

**Authors:** Yongjian Liu, Tianyi Li, Yongqiang Deng, Siyang Liu, Dong Zhang, Hanping Li, Xiaolin Wang, Lei Jia, Jingwan Han, Zhuchun Bei, Yusen Zhou, Lin Li, Jingyun Li

**Author notes:** These authors contributed equally (Yongjian Liu, Tianyi Li, Yongqiang Deng). Corresponding author: Jingyun Li, PhD, State Key Laboratory of Pathogen and Biosecurity, Beijing Institute of Microbiology and Epidemiology, Academy of Military Medical Sciences, Beijing 100071, People’s Republic of China). Lin Li, PhD, State Key Laboratory of Pathogen and Biosecurity, Beijing Institute of Microbiology and Epidemiology, Academy of Military Medical Sciences, Beijing 100071, People’s Republic of China).

## Abstract

At room temperature, SARS-CoV-2 was stable on environmental surfaces and remained viable up to 7 days on smooth surfaces. This virus could survive for several hours in feces and 3-4 days in urine.

---

In the end of 2019, a novel human coronavirus, severe acute respiratory syndrome coronavirus 2 (SARS-CoV-2), emerged and rapidly spread throughout the world. The disease caused by SARS-CoV-2 was named as Coronavirus Disease 2019 (COVID-19) by World Health Organization (WHO). Globally, as of 5 May 2020, there have been 3,525,116 confirmed cases of COVID-19, including 243,540 deaths, reported to WHO. It is an extremely contagious disease with high risk of person-to-person transmission, notably the transmission in hospital and family settings [1, 2]. Prevention and control of SARS-CoV-2 epidemic is based on the knowledge of its transmission route. Although close exposure to respiratory droplets from an infected patient is the main transmission route of SARS-CoV-2, touching contaminated surfaces and objects might also contribute to transmission of this virus. When the droplets produced by a COVID-19 case are too heavy to be airborne, they land on surfaces surrounding the person. A person can be infected with SARS-CoV-2 by touching the contaminated surfaces followed by touching his eyes, nose or mouth[3]. In addition, the evidence for gastrointestinal infection of SARS-CoV-2 and the presence of SARS-CoV-2 RNA in faecal specimens raised a question of a fecal-oral transmission route [4, 5]. Here, we provide a report of our study of the stability of SARS-CoV-2 on various environmental surfaces and in human excreta (feces and urine). The data on the stability of SARS-CoV-2 in different conditions has great importance for our understanding of the transmission route of this virus.

## METHODS

### Viruses and cell line

SARS-CoV-2 strain BetaCoV/Beijing/AMMS01/2020 was originally isolated from the throat swab specimen of a COVID-19 patient. Viral stocks were prepared and titrated on Vero cells as previously described, which were grown in essential Dulbecco’s modified Eagle’s medium (DMEM) containing 2% fetal bovine serum (FBS), 100 U/ml penicillin and 100 μg/ml streptomycin in 5% CO2 at 37°C.

### Stability of SARS-CoV-2 on environmental surfaces

Nine different objects representing a variety of household and hospital situations were collected (stainless steel, plastic, glass, ceramics, paper, cotton clothes, wood, latex gloves, surgical mask). All the materials were cut into small pieces with the area about 1×1cm, which were washed with deionized water and autoclave sterilized at 121°C for 20 min except for latex gloves. The pieces of latex gloves were sterilized with 75% (v/v) ethanol for 30min.

Fifty microliters of virus stock with the infectious titer of 10^6^ 50% tissue culture infectious dose (TCID_50_) per milliliter was deposited on each surface and left at room temperature (25-27°C) with a relative humidity of 35%. At predefined time points (0h, 1h, 2h, 6h, 1d, 2d, 3d, 4d, 5d, 7d), the viruses were recovered by adding 500μl of viral transport medium. The infectivity of residual virus was titrated in quadruplicate on 96-well plates containing 100μl of Vero cells (2×10^5^ cells/ml). The plates were incubated in 5% CO2 at 37°C. On the fifth day, the cytopathic effect (CPE) was observed under a microscope, and the TCID_50_ for each sample at a different time was calculated with Reed-Muench method. All experiments were repeated three times.

### Stability of SARS-CoV-2 in feces and urine

The specimens of feces and urine were collected from three health donors, including two adults and one seven-year-old child. A 10% suspension of each faecal specimen was prepared in PBS (pH, 7.4) as described previously[6]. Faecal suspension and urine samples were filtrated with 0.2 μm filter to remove bacteria. A total of 2.7ml of each filtered faecal suspension and urine sample was inoculated with 0.3ml of virus stock (10^6^ TCID_50_/ml) and left at room temperature for 7days. At desired time points (0h, 1h, 2h, 6h, 1d, 2d, 3d, 4d, 5d, 7d), 50μl of each sample was taken and virus titer was determined with the same method described above.

### The decay of SARS-CoV-2 in experimental conditions

Two-phase linear regression fitting for the log unit TCID_50_/ml against time and estimation of initial and terminal half-lives were performed using R project based on the method for biologic half-life data[7].

## RESULTS

### Stability of SARS-CoV-2 on environmental surfaces

SARS-CoV-2 was stable on plastic, stainless steel, glass, ceramics, wood, latex gloves, and surgical mask, and remained viable for seven days on these seven surfaces. As is shown in Figure 1A, the virus titer declined slowly on these seven surfaces. For example, its TCID^50^/ml decreased from 10^5.83^ at time zero to 10^2.06^ at day 7 on plastic, which was about a 3.8 log10 reduction from the original inoculum. No infectious virus could be recovered from cotton clothes after 4 days and from paper after 5 days (Figure 1A). Rapid loss of infectivity was observed within 1h after incubation on both paper and cotton clothes surfaces. From time zero to 1h, the virus titers decreased from 10^5^ to 10^3.28^ TCID^50^/ml on cotton clothes and from 10^5.56^ to 10^3.44^ TCID^50^/ml on paper, respectively, with an average of about two log10 reduction.

**Figure 1.**
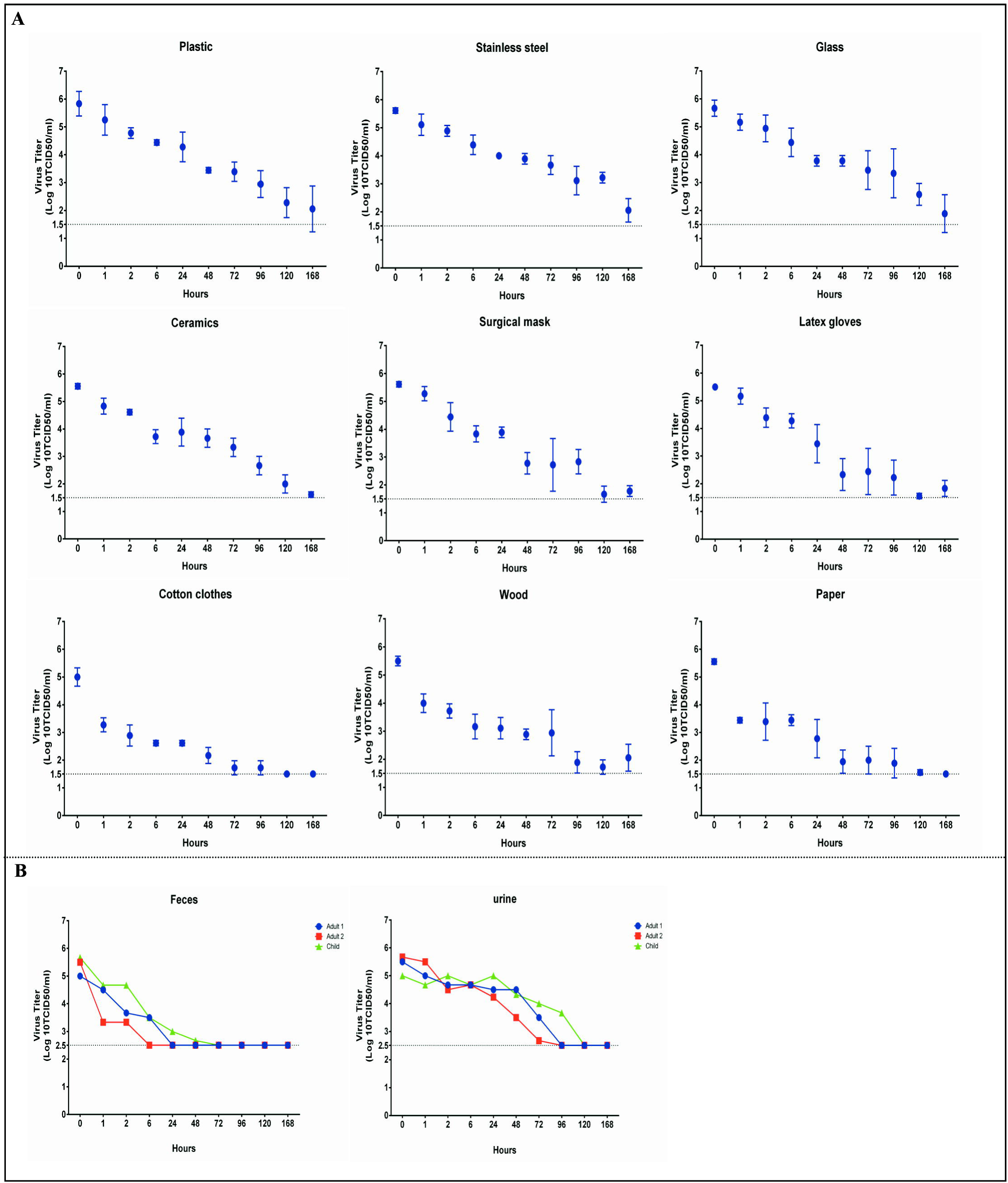
Stability of SARS-CoV-2 on environmental surfaces and in human excreta. (A) Survival time of SARS-CoV-2 on nine surfaces. The Limit of Detection (LOD) for the assays was 10^1.5^ TCID_50_/ml. (B) Survival time of SARS-CoV-2 in three faecal and urine specimens. The LOD for the assays was 10^2.5^ TCID_50_/ml due to cytotoxicity caused by faecal and urine specimens.

### Stability of SARS-CoV-2 in feces and urine

All the three faecal suspension were detected with a PH value of 7.5. Figure 1B shows the duration of SARS-CoV-2 survival in three feces. In the first adult faecal specimen, no viable SARS-CoV-2 was measured after 6 hours, and in the second adult faecal specimen, no virus remained viable after 2 hours. However, the virus survived for 2 days in the child feces, which might indicated the prolonged survival time of SARS-CoV-2 in children’s feces than in adult’s feces. SARS-CoV-2 was more stable in urine than in feces, and infectious virus was detected up to 3 days in two adult urine and 4 days in one child urine.

### The decay of SARS-CoV-2 in experimental conditions

SARS-CoV-2 displayed a two-phase decay in all experimental conditions except for that on paper and in feces (Supplementary Figure). The initial half-lives of SARS-CoV-2 ranged from 0.17h to 0.9h, and rapid loss of infectivity was observed in this phase. However, the terminal half-lives of SARS-CoV-2 with an average of 19.2h were longer than the initial half-lives (Supplementary Table). In the terminal phase, the virus titer decreased slowly. The survival time of SARS-CoV-2 in feces was too short to calculate the half-life, and SARS-CoV-2 presented a one-phase decay after incubation on paper.

## DISCUSSION

The continuously rapid growing of COVID-19 pandemic indicates the great difficulty in controlling and curbing the spread of this disease. As WHO recommended, besides respiratory droplets, fomites such as contaminated objects and surfaces may also serve as transmission sources. A recent study showed evidence that there was extensive environmental contamination by patients with SARS-CoV-2, which suggested the contaminated environment as a potential medium of transmission[8]. Research into the stability of SARS-CoV-2 in different conditions is required urgently. In this study, we provided the information of SARS-CoV-2 stability on nine environmental surfaces, and the data in human excreta (feces and urine) for the first time.

Prior to our study, two research teams had just reported the stability of SARS-CoV-2 on different material surfaces. One study reported by van Doremalen et al found that the viable virus could be detected up to 2-3 days on surfaces of plastic and stainless steel[9]. Chin and colleagues demonstrated the duration of SARS-CoV-2 survival on different surfaces ranged from 2 to 7 days depending on whether the surface is smooth[10]. In comparison with the above two studies, our data displayed a prolonged survival time of this virus on environmental surfaces. In general, the stability of virus in environments was derived from simulate experiments, which were influenced by many factors. The titer of virus stock and the volume of virus inoculation were related with the final results. Compared with van Doremalen’s experiments, we used the same volume of inoculation, but the titer of virus stock was one log unit higher. In Chin’s study, a five microliters of virus stock with the infectious titer of 10^6.8^ TCID_50_/ml was deposited on the surface. Therefore, we highly recommended that a technical specification should be drafted to guide further research into the survival of newly emergent virus.

Fomites such as human excreta is a matter of considerable public concern for the transmission of SARS-CoV-2. Several studies reported the presence of viral RNA in feces of patients in which respiratory samples had switched negative [5, 11]. In this study, we showed that this virus remained viable for several hours in feces and 3-4 days in urine, respectively. With respect to virus isolation, it seems not easy to isolate SARS-CoV-2 from faecal sample, in spite of high virus RNA concentration. Until recently, three SARS-CoV-2 strains were successfully isolated from faecal specimens [12]. The short duration of SARS-CoV-2 survival in feces has practical implications for virus isolation from faecal sample. A reasonable suggestion is that we should shorten the time from sample collection to virus isolation. There was scarce evidence for the presence of viral RNA of SARS-CoV-2 in urine, and no infectious virus has yet been isolated from urine sample. However, isolation of SARS-CoV, closely related to SARS-CoV-2, has indeed been reported. Taken together, the transmission of SARS-CoV-2 by human excreta was plausible.

In conclusion, SARS-CoV-2 displayed stable on environmental surfaces and could survive for several hours in feces and 3-4 days in urine. Effective hand hygiene and adequate disinfection in toilet were recommended to limit the transmission of this virus.

## Data Availability

All the data and reagents presented in this study are available from the corresponding authors upon request

## References

1. Chan JF-W, Yuan S, Kok K-H, et al. A familial cluster of pneumonia associated with the 2019 novel coronavirus indicating person-to-person transmission: a study of a family cluster. LANCET 2020; 395(10223): 514–23.

2. Huang C, Wang Y, Li X, et al. Clinical features of patients infected with 2019 novel coronavirus in Wuhan, China. LANCET 2020; 395(10223): 497–506.

3. Coronavirus disease (COVID-2019): situation report-66. Geneva: World Health Organization, 2020.

4. Xiao F, Tang M, Zheng X, Liu Y, Li X, Shan H. Evidence for Gastrointestinal Infection of SARS-CoV-2. Gastroenterology 2020; (Epub ahead of print).

5. Wu Y, Guo C, Tang L, et al. Prolonged presence of SARS-CoV-2 viral RNA in faecal samples. Lancet Gastroenterol Hepatol 2020; 5(5): 434–5.

6. Lai MYY, Cheng PKC, Lim WWL. Survival of severe acute respiratory syndrome coronavirus. Clin Infect Dis 2005; 41(7): e67-e71.

7. Lee ML, Poon WY, Kingdon HS. A two-phase linear regression model for biologic half-life data. J Lab Clin Med 1990; 115(6): 745–8.

8. Ong SWX, Tan YK, Chia PY, et al. Air, Surface Environmental, and Personal Protective Equipment Contamination by Severe Acute Respiratory Syndrome Coronavirus 2 (SARS-CoV-2) From a Symptomatic Patient. JAMA 2020; (Epub ahead of print).

9. van Doremalen N, Bushmaker T, Morris DH, et al. Aerosol and Surface Stability of SARS-CoV-2 as Compared with SARS-CoV-1. N Engl J Med 2020: NEJMc2004973.

10. Chin AWH, Chu JTS, Perera MRA, et al. Stability of SARS-CoV-2 in different environmental conditions. Lancet Microbe 2020; (Epub ahead of print).

11. An T, Zhen-dong T, Hong-ling W, et al. Detection of Novel Coronavirus by RT-PCR in Stool Specimen from Asymptomatic Child, China. Emerg Infect Dis 2020; (Epub ahead of print).

12. Yao H, Lu X, Chen Q, et al. Patient-derived mutations impact pathogenicity of SARS-CoV-2. medRxiv 2020: 2020.04.14.20060160.

